# A phase Ib/II clinical study to evaluate the safety and efficacy of topical Arnica tincture to treat non-complicated Cutaneous Leishmaniasis in Colombia

**DOI:** 10.1101/2025.06.18.25327211

**Authors:** Sara M. Robledo, Liliana López, Juliana Quintero, Yulied Tabares, Any C. Garcés, Susana Rios, Esteban Soto, Iván D. Vélez, Thomas J. Schmidt

## Abstract

**Background:** Cutaneous leishmaniasis (CL) is caused by *Leishmania* parasites and affects 1.2 million cases annually, mainly in the Americas and the Eastern Mediterranean region. The standard treatment with pentavalent antimonials is often limited by its toxicity, prompting the search for alternative therapies. *Arnica montana* L. (Asteraceae) is a well-known phytotherapeutic plant with anti-inflammatory properties, traditionally used to treat bruises, sprains, distortions, and inflammation caused by insect bites. In our previous work, Arnica tincture (AT) obtained from the flowers showed excellent in vitro activity against *Leishmania* and in vivo activity in a golden hamster model of CL. It also demonstrated high skin permeability and retention in the epidermis without systemic circulation, making it a promising option for topical treatment of CL.

**Methods:** We conducted a randomized, open-label, phase Ib-II clinical trial. Adults with parasitologically confirmed uncomplicated CL were randomly assigned to receive AT topically, 3 times daily, for either 30 or 45 days. We assessed therapeutic response and monitored safety by recording adverse events at each follow-up visit. All adverse events were graded according to the Common Terminology Criteria for Adverse Events (CTCAE), version 5.0.

**Results:** Sixteen subjects were enrolled, with eight assigned to each treatment regimen. Twelve participants completed the 180-day follow-up, all achieved complete lesion healing (100% cure rate). Four participants withdrew their informed consent before or during treatment. The most common adverse events were mild and included erythema, pain, edema, and a burning sensation at the application site. No severe adverse effects were reported.

**Conclusion:** AT showed promising results in treating CL and had a favorable safety profile. Due to the small sample size and lack of comparison with standard therapies, further studies with more robust methodologies are needed to confirm these findings.

Trial registration: NCT05094908

**Author Summary:** Cutaneous leishmaniasis (CL) is a parasitic skin disease affecting over a million people annually, primarily in tropical and subtropical regions. Current treatments rely on pentavalent antimonials, which are often associated with severe side effects and limited accessibility in endemic communities. Seeking safer alternatives, we explored the use of a topical tincture derived from *Arnica montana*, a medicinal plant known for its anti-inflammatory properties. Previous studies have demonstrated the efficacy of this formulation against *Leishmania* parasites in laboratory and animal models, along with high skin retention and low systemic absorption, which are key characteristics for a topical therapeutic. We conducted a phase Ib/II clinical trial in Colombia to assess the safety and therapeutic response of Arnica tincture in adults with parasitologically confirmed, uncomplicated CL (NCT05094908). Participants were randomized to receive treatment for either 30 or 45 days. Of the 16 individuals enrolled, 12 completed the full 180-day follow-up, and all achieved complete lesion healing. No severe adverse effects were reported; only mild, localized reactions such as erythema or burning were observed. Our findings suggest that AT may be a safe and effective topical alternative for treating CL. Although these results are promising, further clinical trials with larger populations and comparison to standard treatments are necessary to confirm its efficacy and potential role in public health strategies.

## Introduction

Tropical Neglected diseases continue to affect hundreds of millions of people worldwide and remain a significant public health concern, particularly in developing countries [1]. Among these diseases, leishmaniasis stands out as a disease that disproportionately impacts populations in tropical and subtropical regions as well as parts of southern Europe. Globally, leishmaniasis affects an estimated 12 million people, with 350 million individuals at risk in nearly 100 countries [2, 3]. Over 90% of new cases of CL are reported in the Americas and the Eastern Mediterranean region, with Afghanistan, Algeria, Brazil, Colombia, Iraq, Pakistan, and Syria collectively accounting for more than 80% of these cases [3, 4].

CL, the most common clinical manifestation, is caused by infection with protozoan parasites of the genus *Leishmania*, transmitted by the bite of infected phlebotomine sandflies [3, 5]. The disease typically begins as a papule at the bite site and can progress over 1 to 3 months to a nodule, an ulcer, or a scaly, wart-like plaque [6]. These disfiguring skin lesions often persist for months or years, leading to significant psychosocial and economic consequences [7].

*Leishmania braziliensis and L. tropica* are the key species responsible for CL in many endemic areas, known for their potential to cause various clinical forms of leishmaniasis; specifically, *L. tropica* is associated with CL in Afro-Eurasia, while *L. braziliensis* is the primary cause of CL and mucocutaneous leishmaniasis in the Americas [2]. Both species are also important for the severity of the disease they cause [2] and their low response to current treatments [8].

For nearly 80 years, first-line treatment for CL typically involves pentavalent antimonials. Despite their efficacy, these drugs pose significant challenges, including parenteral administration, high costs, and notable side effects such as nausea, vomiting, muscle and abdominal pain, cardiac complications, elevated hepatic aminotransferase levels, and chemical pancreatitis. Treatment adherence is often hindered by its lengthy duration (several weeks) and limited availability [4, 9]. Second-line agents such as amphotericin B and pentamidine also pose challenges due to their toxicity profiles and the need for parenteral administration [10]. Miltefosine has demonstrated effectiveness but is associated with teratogenicity and other adverse effects that limit its use [11, 12]. Moreover, access to these drugs can be limited in rural or low-resource settings, further compounding the challenge of disease control.

Given these limitations, the exploration of safe, effective, and locally applicable treatments for CL is crucial. Recently, local treatment has been recommended for patients with localized CL who meet specific criteria (e.g., 1 to 3 lesions in non-critical areas as head and periarticular regions, each lesion <900 mm², and no immunosuppression), including intralesional antimonials or thermotherapy when feasible [4]. However, there remains an urgent need for additional therapeutic options, particularly those derived from plants and applied topically.

*Arnica montana* L. (Asteraceae) is a medicinal plant with a long European tradition. Flower heads are commonly used to treat sprains, bruises, joint complaints, and local muscular pain, as well as superficial inflammation of the skin, such as that caused by insect bites [13, 14, 15]. The plant and its preparations are well-documented to have anti-inflammatory activity [14, 15]. Arnica tincture (AT) is an ethanolic solution prepared from the flowerheads of *A. montana,* according to the European Pharmacopoeia (Ph. Eur.). It is widely used as a traditional herbal medicinal product in Europe for the treatment of external musculoskeletal disorders and superficial inflammatory conditions [16].

Arnica flowers and the AT produced from them contain a mixture of various sesquiterpene lactones (STLs), comprising esters of helenalin and 11a, 13-dihydro helenalin [14, 15, 16]. Such STLs are considered the main active principles behind the well-documented therapeutic effects. Importantly, in addition to their anti-inflammatory properties, our preclinical work has demonstrated that Arnica STLs also exhibit potent activity against trypanosomatid pathogens, including *Trypanosoma* and *Leishmania* species (see the literature cited in [17]. In addition, we previously demonstrated that AT had superior curative efficacy compared to the standard drug glucantime in golden hamsters with experimental CL caused by *L. braziliensis* and *L. tropica* [17, 18]. Furthermore, recent permeability studies have shown that Arnica STLs are rapidly absorbed through porcine and human skin, with most STL (97%) accumulating in the epidermis and binding irreversibly to skin proteins, minimizing systemic exposure and making AT a safer and well-suited option for localized treatments [19].

AT is legally authorized for topical use in several European countries and is included in Colombia vademecum of medicinal plants [20]. These properties and regulatory status, combined with its known anti-inflammatory and wound-healing activities, provide a strong rationale for its investigation as a topical treatment for CL. Therefore, this study aimed to evaluate the safety, tolerance, and preliminary efficacy of AT in patients with uncomplicated CL.

## Methods

### Study design

This is an open-label, randomized (1:1) two-armed phase Ib/II study to evaluate the safety, tolerance, and therapeutic response of two regimens of AT. The protocol was registered at clinicaltrials.gov with the code NCT05094908 (https://clinicaltrials.gov/search?term=NCT05094908). Subjects were enrolled at Grupo de Investigación Clínica PECET (GIC-PECET) in Medellín, Colombia.

### Population

Inclusion/exclusion criteria: Subjects who met the following criteria were included in the study: males and females, aged 18 to 65 years old, with a confirmed parasitological diagnosis of CL, subjects had no more than fourth nodule or ulcerative lesions, each measuring maximum 4 cm (larger diameter), not located on the ear, face, near mucosal membranes, joints, or any area deemed challenging for topical application of the study drug by the principal investigator; the lesion did not exceed an evolution of more than 4 months according to the subject history. Subjects were excluded for the following reasons: females with a positive serum pregnancy test, breast-feeding or of childbearing potential who do not agree to take appropriate contraception during the treatment period and until day 45 after the onset of treatment; additional exclusion criteria included a history of clinically significant medical conditions, as determined by medical history or laboratory results; prior use of antileishmanial drugs within the past 8 weeks; or abnormal baseline laboratory values, defined as hemoglobin <10 g/dL, serum creatinine above the normal range, or ALT/AST levels exceeding three times the upper limit of normal.

The diagnosis of CL was confirmed by at least one of the following methods: microscopic identification of amastigotes in stained lesion tissue, detection of motile promastigotes in aspirate cultures, or identification of *Leishmania* DNA by PCR using previously published protocols [20, 21]. *Leishmania* species were identified using the PCR-RFLP method [21, 22].

Patients who did not meet the eligibility criteria or chose not to participate were provided with standard treatment as outlined by the Colombian Ministry of Health guidelines [13].

### Product under research

The product under investigation is the commercial phytotherapeutic product Tinctura Arnicae e flor. Ph.Eur. Gehrlicher 100 mL manufactured by Gehrlicher Pharmazeutische Extrakte GmbH (Eurasburg, Germany) (https://gehrlicher.de/en/home-2/). The preparation is a 70% hydroethanolic tincture prepared according to the European Pharmacopoeia [16] from the flowers of *Arnica montana* L. The minimum content of STLs, according to Ph. Eur., is 0.04 % m/m, determined as Dihydrohelenalin tiglate. The content of the product batch (Ch.-B. 5866) used in this study was determined by the manufacturer to be 0.044% according to the Ph. Eur. method based on UV detection. A total content of 0.048% was measured by the authors using the LC-MS method to quantify individual STLs published in [19].

### Interventions

Participants received AT for 30 (regimen 1) or 45 days (regimen 2). Applications were performed three times daily at approximately 8-hour intervals (morning, midday, and evening), totaling 90 or 135 applications for regimens 1 and 2, respectively. Approximately 0.5 mL of the tincture was applied per lesion at each application. The tincture was applied topically with a gloved finger to cover the entire ulcerated and indurated area and rubbed gently into the lesion. Before the first application, each day, lesions were cleaned with soap, water, and sterile 0.9% saline and debrided. No other topical treatments, antibiotics, corticosteroids, or cosmetic products were allowed on the lesions during the treatment period. For general hygiene, patients were allowed to use mild, fragrance-free cleansers on intact skin but not on the lesions themselves.

The lesion was covered and left undisturbed until the next application was made. At each subsequent application of the tincture, the previous application and dressing were removed. Study staff members applied the tincture to lesions on day 1 (D1), and then, each participant applied the tincture through D30 or D45, respectively. The application continued until D30 or D45, even if the lesion had obtained 100% re-epithelialization prior to the end of treatment.

### Follow-up and outcomes

Subjects were evaluated at enrollment (Day 1 - D1), at mid-treatment, had a telephone follow-up during treatment (D15/D22±2 days), at the end of treatment (D30/D45±2 days), and then on post-treatment days (PTD): PTD45±5, PTD90±14, and PTD180-14+28. An optional follow-up visit occurs on PTD60±5.

The participant kept a record of the study medication daily application (3 times a day) to document the application of the study medication.

The response to treatment was evaluated clinically. The following definitions were used for each lesion:

- Initial cure: complete re-epithelialization of all ulcers and complete disappearance of the induration at PTD90.
- Final Cure: Cure occurred on PTD90, plus the absence of relapses at PTD180.
- Relapse: a lesion that achieved 100% re-epithelialization by PTD90 that subsequently reopened by PTD180.
- Reactivation: non-healing or worsening of a clinically improving lesion.
- Failure: re-epithelialization of the lesion <50% at PTD45 or <100% re-epithelialization at PTD90, or relapse of the lesion at any time between PTD90 and PTD180, or an increase of ≥100% in ulcer area as compared to baseline, at any time before PTD90.

Subjects who met the failure criteria or chose to withdraw from the study for any reason received rescue therapy-free treatment with meglumine antimoniate, administered by intralesional injection, three to five mL covering the entire ulcer, to be done weekly, with a total of 3 to 5 injections depending on the clinical response, or oral miltefosine at a dose of 1.5 to 2.5 mg/kg/day, with a maximum dose of 150 mg per day, for 28 days, as recommended by the PAHO and the Colombian Ministry of Health [4, 23].

### Study procedures

Clinical evaluation was conducted at enrollment, during treatment, at the end of treatment, and at PTD45, PTD90, and PTD180. All lesions were measured using standard digital calipers and photographed during each visit. Measurements were taken after cleaning the lesion and removing any crust. Using a calibrated electronic caliper, the ulcer dimensions were recorded in two perpendicular directions. The ulcer area was calculated by multiplying the primary diameter (length) by the minor diameter (width).

The percentage of lesion re-epithelialization was determined by comparing the ulcer size at baseline to its size at follow-up visits.

To monitor potential drug-related toxicity, aspartate aminotransferase (AST), alanine aminotransferase (ALT), and creatinine were measured during enrollment and at the end of treatment (D30 or D45). Patients presenting with abnormal laboratory values were monitored until they normalized.

### Adverse Events Monitoring and Grading

Adverse events (AEs) were actively monitored and recorded during each scheduled study visit (at baseline, mid-treatment, end-of-treatment, and follow-up visits), as well as during telephone follow-ups. Participants were also asked to document any symptoms in a daily treatment diary. All adverse events were graded and classified according to the Common Terminology Criteria for Adverse Events (CTCAE) version 5.0 [24].

### Sample size

Sixteen subjects, eight per treatment group (a number chosen for convenience), comparable through rigorous selection and application of eligibility criteria, will provide initial evidence of AT efficacy in treating uncomplicated CL.

### Randomization process

A computer-generated randomization code using a 1:1 allocation ratio was used to assign subjects to either treatment arm.

The randomization sequence was implemented using numbered, sealed, opaque envelopes. The pharmacist who guarded these envelopes was the only person authorized to open them and indicate the assigned treatment group.

### Statistical Analysis

Data were verified through double entry prior to analysis. The t-test or Mann–Whitney test was used to compare quantitative data based on their distribution. Differences in proportions were assessed using the χ² test or Fisher exact test (when the expected frequency in any cell was <5). The odds ratio and proportion of definitive cure by treatment arm (primary outcome) were calculated. Additionally, the frequency of adverse events by the treatment group was estimated, and these events were compared based on severity, intensity, and their relationship with the study intervention. Differences in cure proportions at PTD45 and PTD90 (secondary outcomes) were also calculated.

Analyses were conducted using intention-to-treat (ITT) and per-protocol (PP) methods using IBM SPSS Statistics para Windows (Version 29). A p-value of < 0.05 was considered statistically significant. The PP analysis included only those patients who completed follow-up visits and received at least 90% of the prescribed tincture dose.

### Ethical Approval and Regulation

The protocol for this study was designed based on the general ethical principles contemplated in the Declaration of Helsinki and the guidelines of the ICH (International Committee for Harmonization) on Good Clinical Practice. The Research Ethics Committee of CLINISALUD del Sur S.A.S. approved this study by act CEI-0101-09-2021. The trial was conducted following Good Clinical Practice standards.

## Results

### Participant characteristics

Participants were enrolled from May 2023 to September 2024. Sixteen participants were randomly assigned to one of the two treatment arms (AT 30 days or AT 45 days). In total, 12 subjects (n=6 ‘30-day scheme’ and n=6 ‘45-day scheme’) completed the study follow-up. None relapse or reactivation occurred. The description of treatment outcomes, voluntary withdrawals, and loss to follow-up are shown in Figure 1.

**Figure 1.**
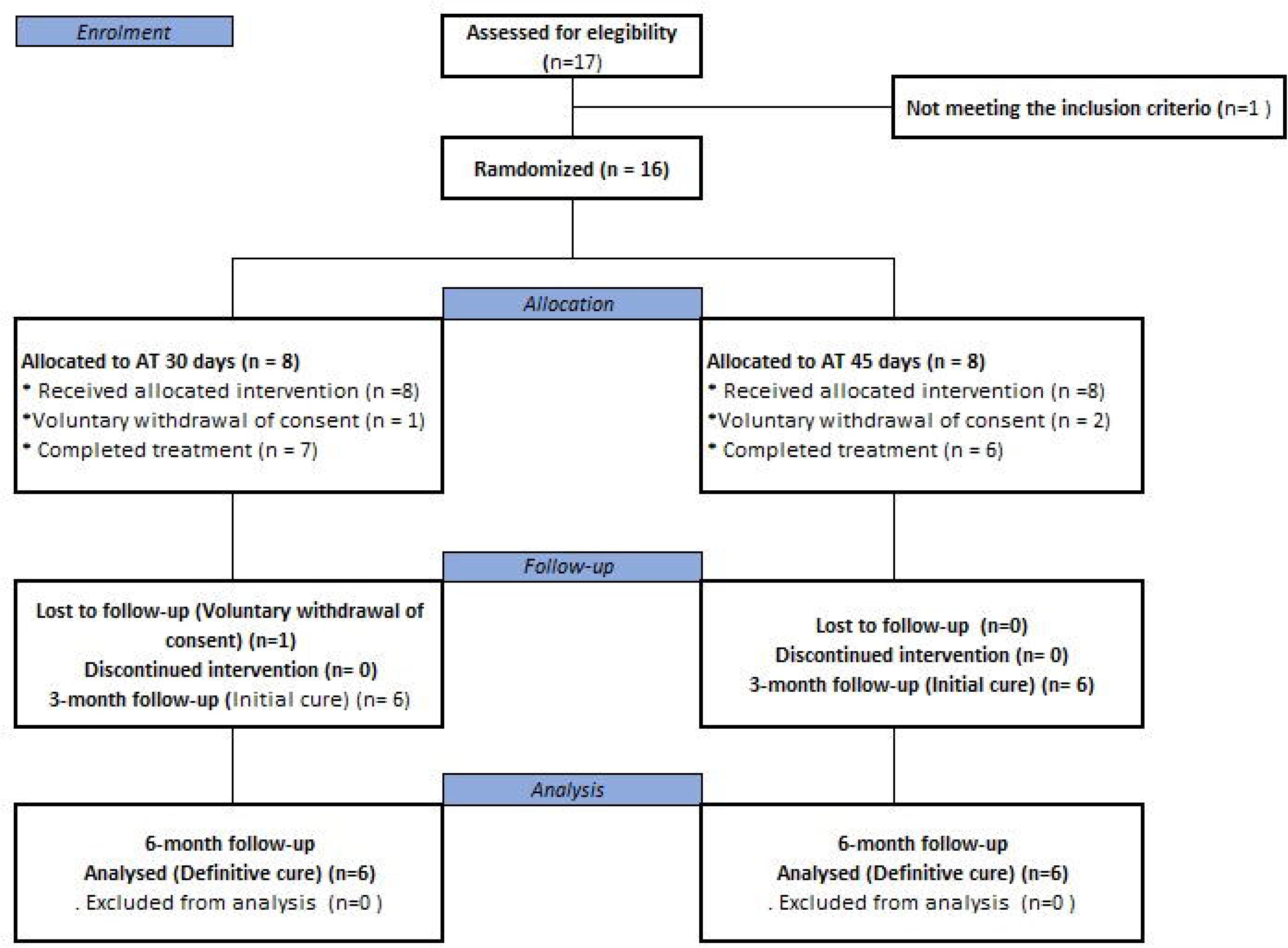
Enrollment and follow-up of participants.

Table 1 presents the baseline and disease descriptions by treatment group. Randomization successfully assigned subjects with characteristics similar to those of the two treatment groups.

**Table 1.**
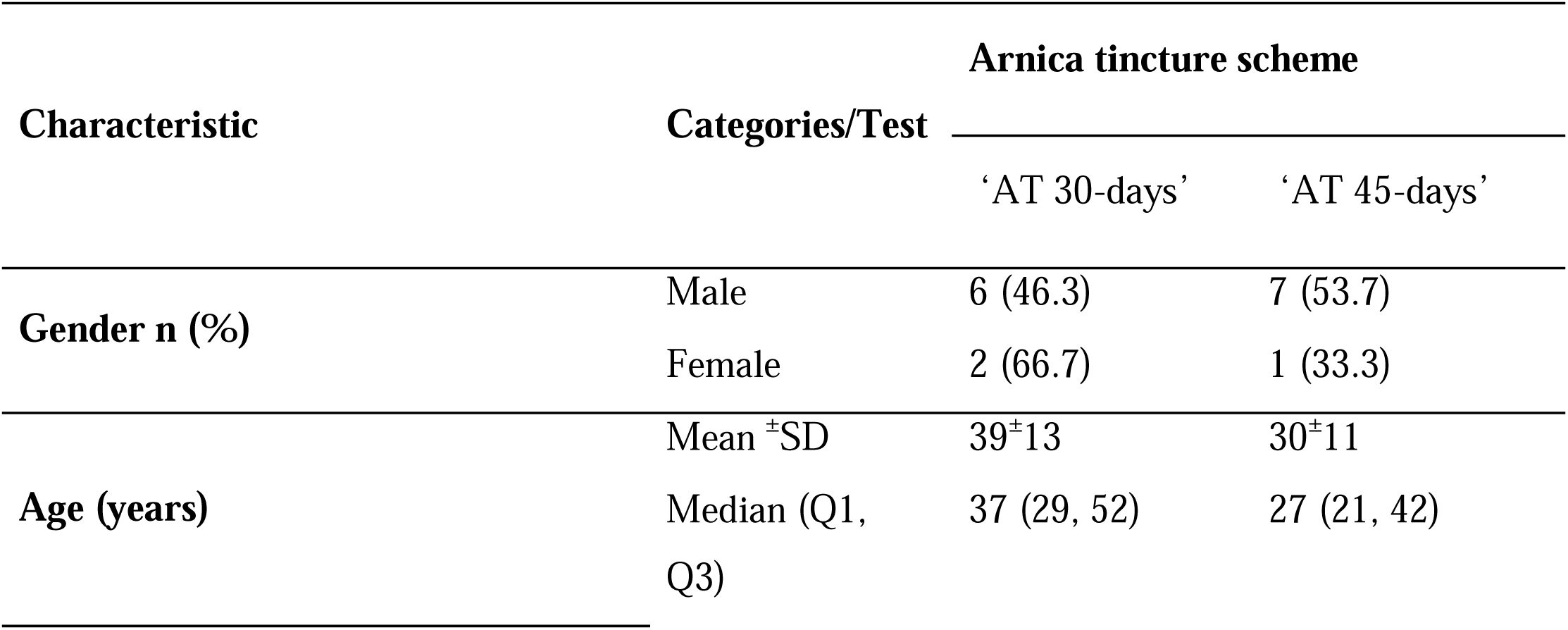
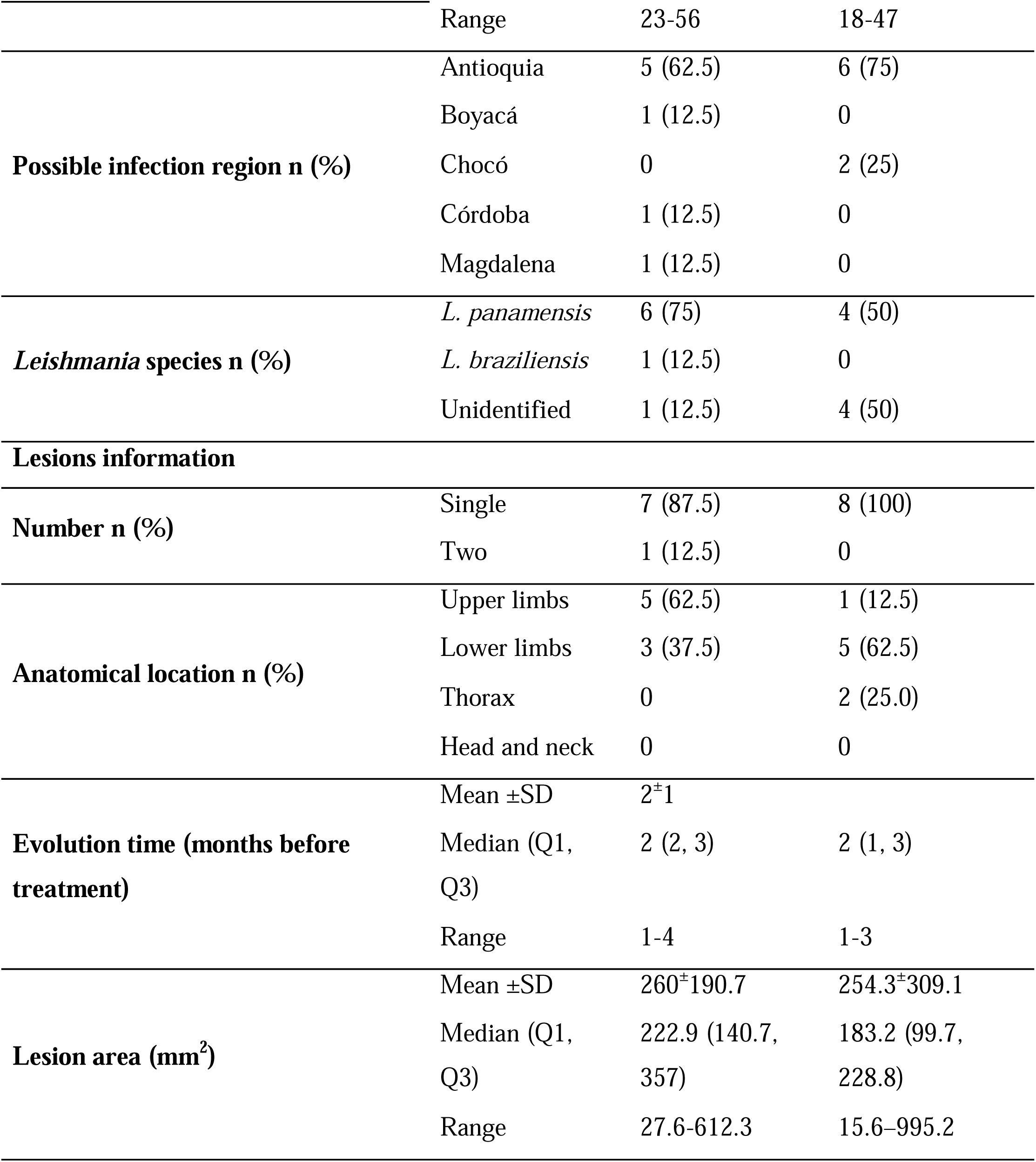
Baseline participant characteristics and disease description.

The entire study population was mixed race, with no history of infection with Leishmaniasis and all had ulcerated lesions. Except for adenopathies in the area of the lesion (n = 1), the physical examination was regular in all participants. At screening, liver (aspartate aminotransferase and alanine aminotransferase) function tests were regular; one participant had a non-clinically significant abnormal creatinine level (renal function).

### Therapeutic Response

Of the 12 participants (n=6, 30-day scheme, and n=6, 30-day scheme) who completed the entire study, including the 180-day follow-up, all achieved complete lesion healing, demonstrating a 100% cure rate (Table 2). The remaining four participants withdrew their informed consent before or during treatment (see Table S1, Supporting Information).

**Table 2.**
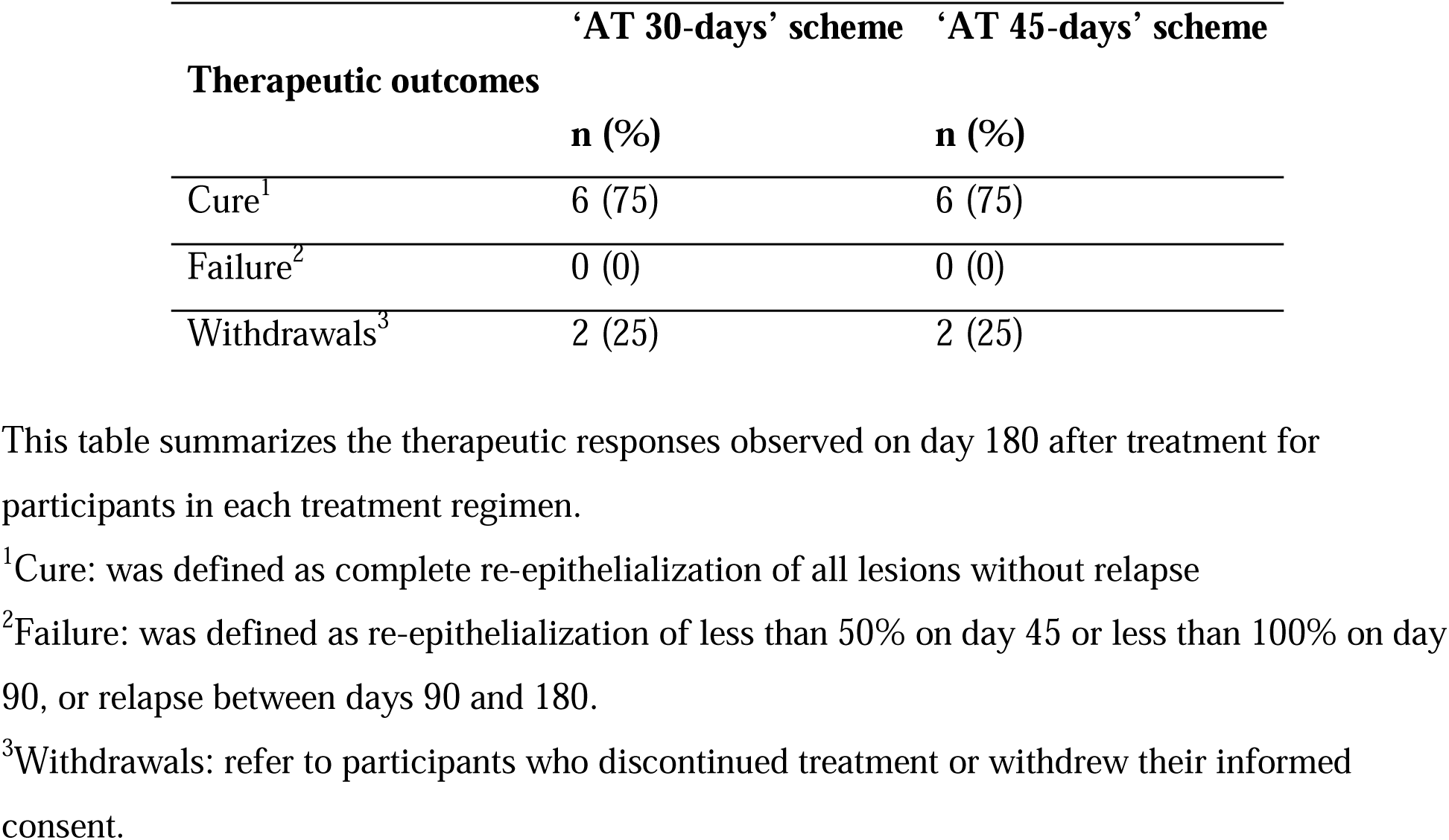
Therapeutic outcomes of participants receiving Arnica tincture for cutaneous leishmaniasis.

| Therapeutic outcomes | 'AT 30-days' scheme | 'AT 45-days' scheme |
| --- | --- | --- |
|  | n (%) | n (%) |
| Cure<sup>1</sup> | 6 (75) | 6 (75) |
| Failure<sup>2</sup> | 0 (0) | 0 (0) |
| Withdrawals<sup>3</sup> | 2 (25) | 2 (25) |

This table summarizes the therapeutic responses observed on day 180 after treatment for participants in each treatment regimen.
<sup>1</sup>Cure: was defined as complete re-epithelialization of all lesions without relapse
<sup>2</sup>Failure: was defined as re-epithelialization of less than 50% on day 45 or less than 100% on day 90, or relapse between days 90 and 180.
<sup>3</sup>Withdrawals: refer to participants who discontinued treatment or withdrew their informed consent.

Figure 2 shows information on the evolution of lesion size during the treatment and post treatment (PT) follow-up; except for 1 participant (45-day scheme), all ulcers showed a significant decrease in area at PT day 45, with healing in 56% (n=9) of them. The underlying numeric data, presented in tabular form, is shown in Table S1 of the Supporting Information.

**Figure 2.**
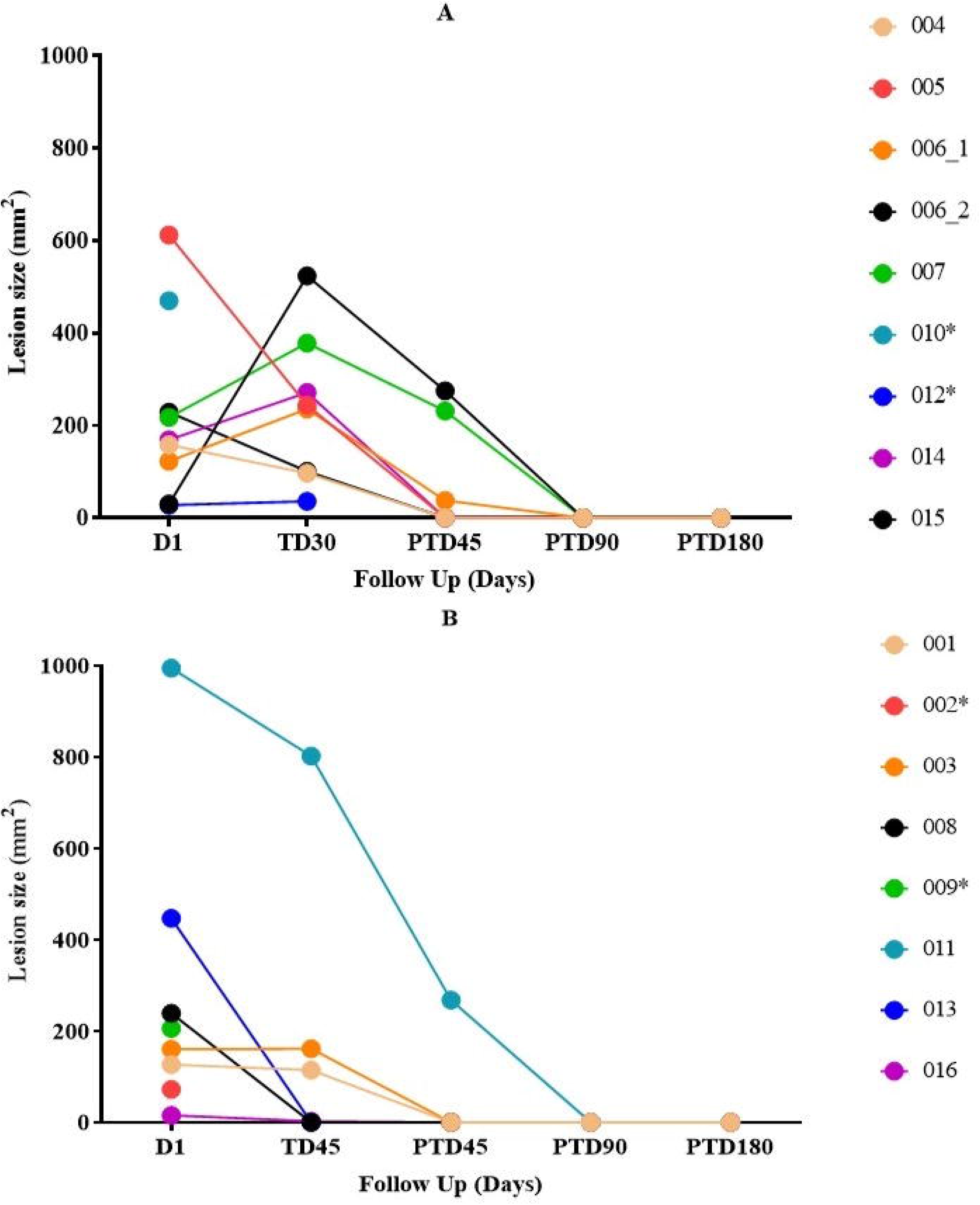
Lesion size evolution during the treatment and post-treatment (PT) follow-up. The figures represent individual lesion trajectories across different subjects. D1 indicates the start of treatment, while TD30 (A) and TD45 (B) denote 30 and 45 days of treatment duration, respectively. PTD45, PTD90, and PTD180 correspond to 45, 90, and 180 days post-treatment, respectively. Each line represents the progression of lesion size for an individual participant. * corresponds to participants who withdrew from the study.

Figure 3 presents photos illustrating the healing progress of a representative patient (Patient 015, as shown in Figure 2). The corresponding photo documentation for all patients is provided in Figures S1 and S2 of the Supporting Information.

**Figure 3.**
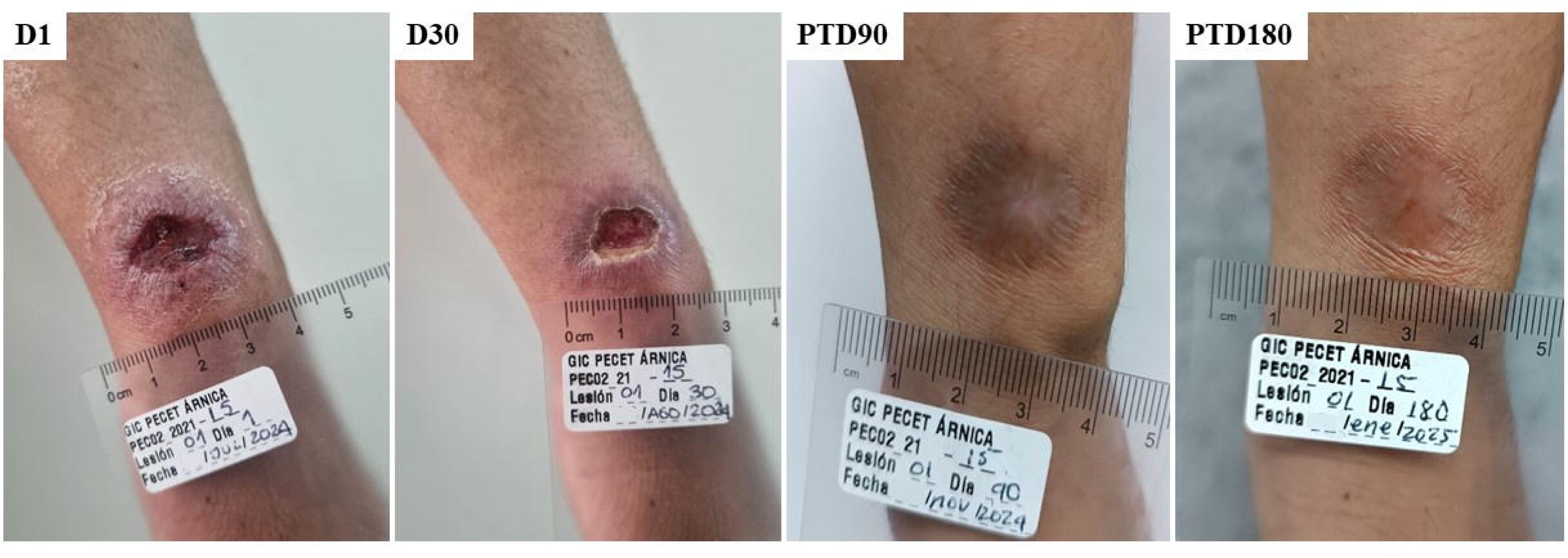
Evolution of the lesion during treatment and post-treatment (PT) follow-up. The figures represent the lesion trajectories for one representative patient across the study. D1 indicates the start of treatment, TD30 is the last day of treatment, and PTD90 and PTD180 correspond to 90 and 180 days post-treatment, respectively.

#### Safety (Adverse events)

Seven subjects in the AT 30-day group and six in the AT 45-day group reported a total of 41 adverse events (AEs) related to tincture use. Erythema, burning sensation, and pruritus were the most common local AEs, all affecting the area around the lesion where the tincture was applied (Table 3). All adverse events were graded according to the CTCAE version 5.0 [24], with the majority being mild to moderate in severity. Seven subjects reported adverse events that were classified as not related to the study drug, including flu-related symptoms (2), bacterial over-infection in the lesion (1), cellulitis (1), contact dermatitis (1), sharps injury (1), and headache (1). There were no serious AEs.

**Table 3.** Incidence of local adverse events (AE) by treatment group, as reported by the study participants.

| Adverse event |  | ‘AT 30-days’ scheme | ‘AT 45-days’ scheme |
| --- | --- | --- | --- |
| Number of AE reported<sup>1</sup> |  | 24 (7 subjects= 88%) | 17 (6 subjects= 75%) |
| Local adverse event | Grade<sup>2</sup> | n (%) | n (%) |
| **Erythema** | Grade 1 / Mild | 4 (80) | 1 (20) |
|  | Grade 2 / Moderate | 0 | 1 (20) |
|  | Grade 3 / Severe | 1 (20) | 2 (60) |
| **Burning sensation** | Grade 1 / Mild | 5 (71.4) | 4 (66.6) |
|  | Grade 2 / Moderate | 2 (28.6) | 1 (16.7) |
|  | Grade 3 / Severe | 0 | 1 (16.7) |
| **Pain** | Grade 1 / Mild | 3 (100) | 1 (50) |
|  | Grade 2 / Moderate | 0 | 1 (50) |
| **Pruritus** | Grade 1 / Mild | 4 (80) | 1 (33) |
|  | Grade 2 / Moderate | 0 | 1 (33) |
|  | Grade 3 / Severe | 1 (20) | 1 (33) |
| **Edema** | Grade 2 / Moderate | 3 (75) | 2 (100) |
|  | Grade 3 / Severe | 1 (25) | 0 |

<sup>1</sup>No difference in adverse event frequencies between groups (p=0.52, chi-square test).
<sup>2</sup>Grading according to Common Terminology Criteria for Adverse Events (CTCAE) v5.0 [24].

During the screening, one participant presented a slight alteration in the creatinine level, which was considered not clinically significant due to the absence of relevant clinical conditions in the participant. The result was attributed to possible dehydration related to the fasting required for sample collection. Laboratory tests of renal and hepatic function of all participants were in the normal range at the end of treatment (see Table S2, Supporting Information).

In terms of adherence, 8 participants (n=5 in the ‘AT 30-days’ scheme and n=3 in the ‘AT 45-days’ scheme) missed at least one dose of AT, but none met the criteria for discontinuation; the reason for the missed doses was forgetfulness. No subject missed a full day’s dosing.

## Discussion

This study was conducted in accordance with the approved protocol and the principles of good clinical practice. All participants voluntarily agreed to participate in the study and received their treatment according to the treatment arm, which they were randomly allocated. The follow-up rate at 6 months was 75%. Of the 12 participants who completed the 180-day follow-up, all achieved complete lesion healing, demonstrating a 100% cure rate.

This open-label and randomized Phase Ib-II clinical trial demonstrated that AT applied topically to CL lesions achieved a therapeutic response of 100% in patients who completed the 180-day post-treatment follow-up. These results suggest superior efficacy compared to conventional therapies, such as miltefosina [25] or amphotericin B [26], with the added advantage of being a non-invasive topical treatment. This suggests that AT could represent a therapeutic alternative for patients who do not tolerate systemic treatments or who prefer more accessible and less aggressive options.

Concerning the therapeutic regimens evaluated, the finding of no significant differences in clinical outcomes between the 30-day and 45-day groups is important. From the perspective of clinical practice and patient experience, this equivalence in therapeutic response favors the shorter 30-day regimen, which would reduce the total application time by approximately one-third (favoring adherence) [17] and consequently reduce patient exposure to the AT treatment.

The observed adverse events were of low severity, consistent with expectations for a topical treatment [28]. This reinforces the safety profile of AT and its potential to be administered on an outpatient basis with minimal physician supervision. The absence of serious adverse events is particularly relevant, given the significant toxicity associated with current systemic treatments.

Although the exact mechanisms of action of AT in the context of CL were not explored in this study, AT has a long history of traditional use in European herbal medicine for the topical treatment of bruises, sprains, and inflammatory skin conditions [13]. Its pharmacological effects are attributed mainly to the presence of STLs, particularly helenalin and dihydrohelenalin derivatives, which are the main active constituents that have demonstrated anti-inflammatory, antimicrobial, and wound-healing properties [27]. These STLs inhibit the activation of NF-κB and the expression of pro-inflammatory cytokines, thereby modulating the local immune response and reducing tissue damage [28]. Additionally, AT has been shown to promote re-epithelialization by stimulating cell migration and proliferation, processes essential for wound closure and healing [18]. While direct studies on the Arnica effect in CL are limited, its known anti-inflammatory and wound-healing effects, along with in vitro and in vivo evidence of antiprotozoal, particularly anti-leishmanial, activity [17, 18], provide a plausible rationale for its use in this context. These established pharmacological activities may explain the dual effect observed in this trial and could explain the high percentage of healing observed. Compared with conventional antileishmanial drugs, which primarily act on the parasite, AT could offer the additional advantage of modulating the local inflammatory response and creating an environment favorable to healing.

Our findings align with the growing interest in natural or plant-based treatments for CL. While most research on herbal products remains preclinical, a recent clinical study by Aghaei et al. (2024) [29] evaluated an herbal ointment containing Aloe vera, Brazembel, Nigella sativa, Propolis, Lavender, and Olive oil in 60 Iranian patients with CL. This study demonstrated a significant reduction in lesion size in the herbal ointment group compared to the control group receiving intralesional glucantime, suggesting that multi-component plant-based preparations may provide an effective alternative to conventional treatments. Notably, while Shirvan herbal ointment showed promise, it remains an investigational product without regulatory approval. In contrast, AT is already approved as a traditional herbal medicinal product by the European Medicines Agency (EMA) and is commercially available in other countries as a phytotherapeutic or cosmetic product, facilitating broader access and implementation compared to other natural agents still under experimental investigation.

The small sample size of this study was intentionally chosen, as this Phase Ib-II trial was designed as a pilot study to assess the initial safety, tolerability, and early efficacy signals of AT. The small sample size was not due to a low disease burden, as CL is endemic in the study area. Instead, it was influenced by practical and ethical considerations: recruitment was limited to a single clinical site with a rigorous eligibility process, and patients ineligible for the study received immediate standard care. Notably, a placebo group was not included because in clinical practice, thousands of patients have shown that alcohol alone, used as the only excipient in Arnica tincture has no leishmanicidal activity and does not cure cutaneous leishmaniasis, making the use of placebo ethically unacceptable. Furthermore, the primary aim of this study was to compare two different treatment regimens of AT to evaluate differences in safety and efficacy rather than to deprive a control group of a potentially beneficial treatment. The primary focus was on generating preliminary evidence to support future, larger studies.

Despite the small sample size and the lack of a placebo control group, the findings are relevant for public health because they provide an early indication of the potential utility of AT as a safe, effective, and locally applicable treatment. These data are valuable for the design of larger and more definitive randomized controlled trials.

Finally, the ease of topical application and favorable safety profile make AT especially attractive for rural settings or communities with limited access to specialized health services. As a plant-derived preparation, it also holds the potential for more sustainable and affordable production compared to synthetic drugs.

## Conclusion

AT demonstrated promising results in treating CL and exhibited a favorable safety profile. Its topical, non-invasive administration and potential dual action (anti-leishmanial and anti-inflammatory) suggest that it could be an attractive alternative for patients who do not tolerate systemic therapies or for those in remote areas with limited access to specialized care.

In terms of public health relevance, AT could help reduce the treatment burden for CL, particularly in rural or resource-limited settings. As a locally applicable, plant-derived product, it may offer an affordable and accessible option with a lower risk of systemic adverse effects compared to current first-line therapies such as pentavalent antimonials, amphotericin B, or miltefosine. Importantly, scaling up production and distribution would be supported by the fact that AT is already approved by the European Medicines Agency (EMA) as a traditional herbal medicinal product and is commercially available in other countries as a homeopathic or cosmetic product. This established regulatory and commercial framework facilitates broader access while ensuring consistent quality and safety.

Future studies should include larger sample sizes, direct comparisons with standard treatments and cost-effectiveness analyses to integrate AT into existing treatment guidelines for CL. Such studies are already underway.

## Limitations of the study

This study employed an open-label (non-blinded) design, meaning that both participants and investigators were aware of the assigned treatment. This approach was necessary due to the inherent difference in the duration of the two treatment regimens (30 vs. 45 days), which was known to both patients and study staff during the mid-treatment and end-of-treatment visits. Consequently, blinding was not feasible in this context.

Notably, a placebo group was not included because in clinical practice, thousands of patients have shown that alcohol alone, used as the only excipient in Arnica tincture has no leishmanicidal activity and does not cure cutaneous leishmaniasis, making the use of placebo ethically unacceptable. Furthermore, the primary aim of this study was to compare two different treatment regimens of AT to evaluate differences in safety and efficacy rather than depriving a control group of a potentially beneficial treatment.

The sample size (16 enrolled, 12 completed) was small and chosen for feasibility as an exploratory Phase Ib-II study rather than based on formal power calculations for efficacy. The small sample size reflects the pilot nature of the trial, strict eligibility criteria, and ethical considerations in this population, not a low disease burden. Although this limits the statistical inference and generalizability of the findings, the results provide important preliminary data to guide future, larger-scale clinical trials.

The PCR-HPLC was unable to identify some Leishmania strains in this study. This limitation is likely due to genetic variability beyond the scope of the current reference database. To address this, future work will incorporate direct sequencing of target regions to enable the identification of novel or divergent strains, thereby enhancing the robustness of the reference database.

Future research should include a larger number of participants and more severe cases of CL to confirm these findings and establish the potential role of AT within clinical guidelines.

## Supporting information

**S1 Table. Evolution of lesions in each participant during the study.**

**Table S2. Serum Metabolite Levels (AST, ALT, and Creatinine) Before and After Treatment in Study Participants.**

**S1 Figure. Photographies of the evolution of lesions.** The figures represent individual lesion trajectories across different subjects. D1 indicates the start of treatment, while TD30 (A) and TD45 (B) denote 30 or 45 days of treatment duration, respectively. PTD45, PTD90, and PTD180 correspond to 45, 90, and 180 days post-treatment, respectively. Each line represents the progression of lesion size for an individual participant. “*” corresponds to participants who withdrew from the study.

## Data Availability

The data obtained in this study are contained within the article. However, the underlying raw data are available on request from the corresponding author.

## Data Availability

All data generated or analyzed during this study are included in this published article and its supplementary information files and are available from the corresponding author upon reasonable request.

## Acknowledgments

The authors are very grateful to Wilhelm Doerenkamp-Foundation, Chur, Switzerland, for financial support of this study in the form of a NATVANTAGE GRANT. We also thank Gehrlicher Pharmazeutische Extrakte GmbH, Eurasburg, Germany, for supporting this study by donating the Arnica tincture. We thank Dr. Franziska Jürgens, Münster, Germany, for developing the analytical method to determine the exact STL content of Arnica tincture and Lizanne Schäfer, Münster, for analyzing the tincture batch used in this study. This study is part of the Research Network on Natural Products against Neglected Diseases (ResNet NPND; see www.resnetnpnd.org).

## Author Contributions

**Conceptualization:** Sara M. Robedo and and Thomas J. Schmidt.

**Data curation:** Juliana Quintero, Yulied Tabares, Any C. Garcés

**Formal analysis:** Liliana López, Iván D. Vélez and Sara M. Robledo

**Funding adquisition:** Thomas J. Schmidt and Sara M. Robedo

**Investigation:** Juliana Quintero, Yulied Tabares, Any C. Garcés, Sara M. Robledo, Susana Ríos and Esteban Soto.

**Methodology**: Liliana López, Iván D. Vélez, Sara M. Robledo.

**Project administration:** Thomas J. Schmidt

**Resources:** Thomas J. Schmidt and Sara M. Robledo.

**Supervision:** Thomas J. Schmidt.

**Visualization:** Liliana López, Sara M. Robledo, and Thomas J. Schmidt.

**Writing—original draft preparation:** Sara M. Robledo, Thomas J. Schmidt, Liliana López and Juliana Quintero.

**Writing—review and editing:** Thomas J. Schmidt, Liliana López, Iván D. Vélez and Sara M. Robledo

All authors have read and agreed to the published version of the manuscript.

Reporting of the trial was prepared following the CONSORT guidelines [30,31].

