## Supplemental Table 1 and 2 for "A phase Ib/II clinical study to evaluate the safety and efficacy of topical Arnica tincture to treat non-complicated Cutaneous Leishmaniasis in Colombia"

**SUPPORTING INFORMATION**

**Table S1. Evolution of lesions in each participant during the study**

| **Code** | **Lesion type** | **N° lesion** | **AT scheme^a^** | **Lesion area (mm^2^)** | | | | | | **Outcome** |
| --- | --- | --- | --- | --- | --- | --- | --- | --- | --- | --- |
|  |  |  |  | **D1** | **D30 or D45** | **PTD45** | **PTD60^b^** | **PTD90** | **PTD180** |  |
| PEC02-21_001 | Ulcer | 1 | 45 | 126.83 | 114.62 | 0 | - | 0 | 0 | Definitive Cure |
| PEC02-21_002 | Ulcer | 1 | 45 | 72.48 | NA^c^ | NA | NA | NA | NA | Withdrawal^d^ |
| PEC02-21_003 | Ulcer | 1 | 45 | 160.34 | 161.35 | 0 | - | 0 | 0 | Definitive Cure |
| PEC02-21_004 | Ulcer | 1 | 30 | 158.26 | 96.51 | 0 | - | 0 | 0 | Definitive cure |
| PEC02-21_005 | Ulcer | 1 | 30 | 612.16 | 243.94 | 0 | - | 0 | 0 | Definitive cure |
| PEC02-21_006 | Ulcer | 1 | 30 | 122.47 | 235.65 | 37.66 | - | 0 | 0 | Definitive cure |
|  | Ulcer | 2 |  | 29.69 | 523.70 | 274.80 | - | 0 | 0 | Definitive cure |
| PEC02-21_007 | Ulcer | 1 | 30 | 217.36 | 378.37 | 231.47 | 52,40 | 0 | 0 | Definitive cure |
| PEC02-21_008 | Ulcer | 1 | 45 | 239.18 | 0,00 | 0 | - | ND^e^ | ND | Cure |
| PECO2-21_009 | Ulcer | 1 | 45 | 206.06 | NA | NA | NA | NA | NA | Withdrawal |
| PEC02-21_010 | Ulcer | 1 | 30 | 470.02 | NA | NA | NA | NA | NA | Withdrawal |
| PEC02-21_011 | Ulcer | 1 | 45 | 995.24 | 802.49 | 268.13 | - | 0 | 0 | Definitive cure |
| PEC02-21_012 | Ulcer | 1 | 30 | 27.55 | 36.10 | NA | NA | NA | NA | Withdrawal |
| PEC02-21_013 | Ulcer | 1 | 45 | 447.28 | 0 | 0 | - | 0 | 0 | Definitive cure |
| PEC02-21_014 | Ulcer | 1 | 30 | 168.79 | 271.13 | 0 | - | 0 | 0 | Definitive cure |
| PEC02-21_015 | Ulcer | 1 | 30 | 228.55 | 100.60 | 0 | - | 0 | 0 | Definitive cure |
| PEC02-21_016 | Ulcer | 1 | 45 | 15.57 | 2.36 | 0 | - | 0 | 0 | Definitive cure |

^a^ AT: Arnica Tincture. ^b^Optional visit. ^c^NA: Not applicable. ^d^Participants retired the informed consent or they were retired because they were also treated with glucantime or miltefosine. ^e^Non-compliance in follow-up visits.

**Table S2. Serum Metabolite Levels (AST, ALT, and Creatinine) Before and After Treatment in Study Participants**

| **Participant ID** | **Screening** | | | | | | **End of the treatment** | | | | | |
| --- | --- | --- | --- | --- | --- | --- | --- | --- | --- | --- | --- | --- |
|  | **AST (U/L)** | **Reference value (U/L)** | **ALT (U/L)** | **Reference value (U/L)** | **Creatinine (mg/dl)** | **Reference value mg/dL** | **AST (U/L)** | **Reference value (U/L)** | **ALT (U/L)** | **Reference value (U/L)** | **Creatinine (mg/dl)** | **Reference value mg/dL** |
| PEC02-21_001 | 19 | 0 - 34 | 27 | 0 - 55 | 0.62 | 0.55 – 1.02 | 14 | 0 - 34 | 10 | 0 - 55 | 0.54 | 0.55 – 1.02 |
| PEC02-21_002 | 21 | 5 - 34 | 22 | 0 - 55 | 0.86 | 0,73 -1.18 | NA | NA | NA | NA | NA | NA |
| PEC02-21_003 | 13 | 14 - 35 | 12 | 8 - 22 | 0.78 | 0.73 -1.18 | 17,71 | 14 - 35 | 14,1 | 8 - 22 | 0.89 | 0.73 -1.18 |
| PEC02-21_004 | 18 | 5 - 34 | 22 | 0 - 55 | 0.8 | 0,73 -1.18 | 25 | 5 - 34 | 39 | 0 - 55 | 0.77 | 0.73 -1.18 |
| PEC02-21_005 | 22 | 5 - 34 | 19 | 0 - 55 | 0.91 | 0.55 – 1.02 | 22 | 11 -34 | 16 | 0 - 45 | 0.88 | 0.73 -1.18 |
| PEC02-21_006 | 32 | 11 - 34 | 64 | 0 - 45 | 0.74 | 0.73 -1.18 | 36 | 11 -34 | 64 | 0 - 45 | 1.17 | 0.73 -1.18 |
| PEC02-21_007 | 30 | 11 - 34 | 62 | 0 - 45 | 0.96 | 0.73 -1.18 | 20 | 11 -34 | 38 | 0 - 45 | 0.96 | 0.73 -1.18 |
| PEC02-21_008 | 22 | 11 - 34 | 18 | 0 - 45 | 0.89 | 0.73 -1.18 | 22 | 11 -34 | 18 | 0 - 45 | 0.94 | 0.73 -1.18 |
| PEC02-21_009 | 13 | 14 - 35 | 7 | 8 - 22 | 1.12 | 0.73 -1.18 | NA | NA | NA | NA | NA | NA |
| PEC02-21_010 | 19 | 11 - 34 | 18 | 0 - 45 | 1.03 | 0.73 -1.18 | NA | NA | NA | NA | NA | NA |
| PEC02-21_011 | 31 | 11 - 34 | 44 | 0 - 45 | 0.88 | 0.73 -1.18 | 24 | 11 - 34 | 30 | 0 - 45 | 0.91 | 0.73 -1.18 |
| PEC02-21_012 | 14 | 11 - 34 | 9 | 0 - 34 | 0.72 | 0.55 – 1.02 | 19 | 11 - 34 | 17 | 0 - 34 | 0,69 | 0.55 – 1.02 |
| PEC02-21_013 | 25 | 14 - 35 | 31 | 9 - 24 | 0.93 | 0.73 -1.18 | 31 | 11 - 34 | 28 | 0 - 45 | 0,85 | 0.73 -1.18 |
| PEC02-21_014 | 19 | 11 - 34 | 14 | 0 - 34 | 0.73 | 0.55 – 1.02 | 18 | 11 - 34 | 14 | 0 - 34 | 0,80 | 0.55 – 1.02 |
| PEC02-21_015 | 27 | 11 - 34 | 41 | 0 - 45 | 0.84 | 0.73 -1.18 | 26 | 11 - 34 | 36 | 0 - 45 | 0,76 | 0.73 -1.18 |
| PEC02-21_016 | 19 | 11 - 34 | 12 | 0 - 45 | 1.25 | 0.73 – 1.18 | 18 | 11 - 34 | 15 | 0 - 45 | 1,18 | 0.73 – 1.18 |

AST: Aspartate aminotransferase; ALT: Alanine aminotransferase. Reference values are included for comparison. “NA” indicates values not applicable or not measured for that participant. All metabolite levels are presented in U/L (AST, ALT) or mg/dL (Creatinine).

**Figure S1. Photography of evolution of lesions.**

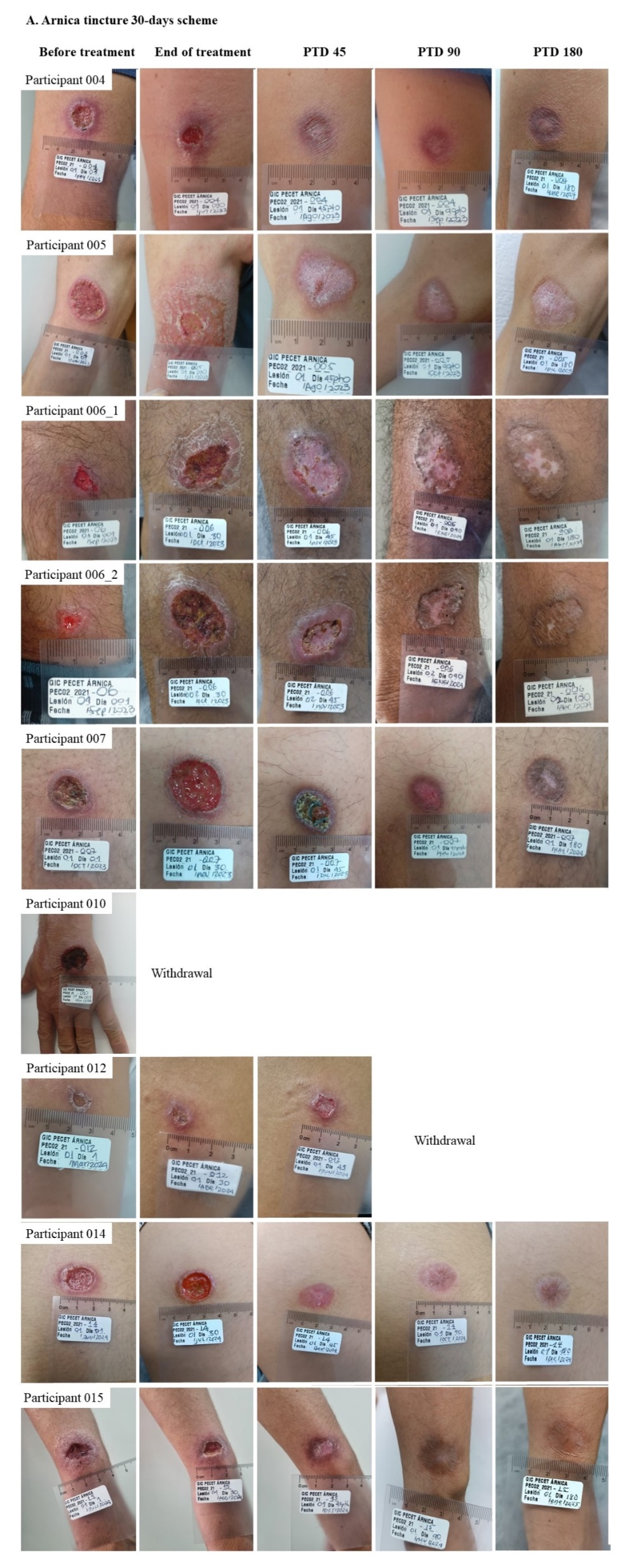

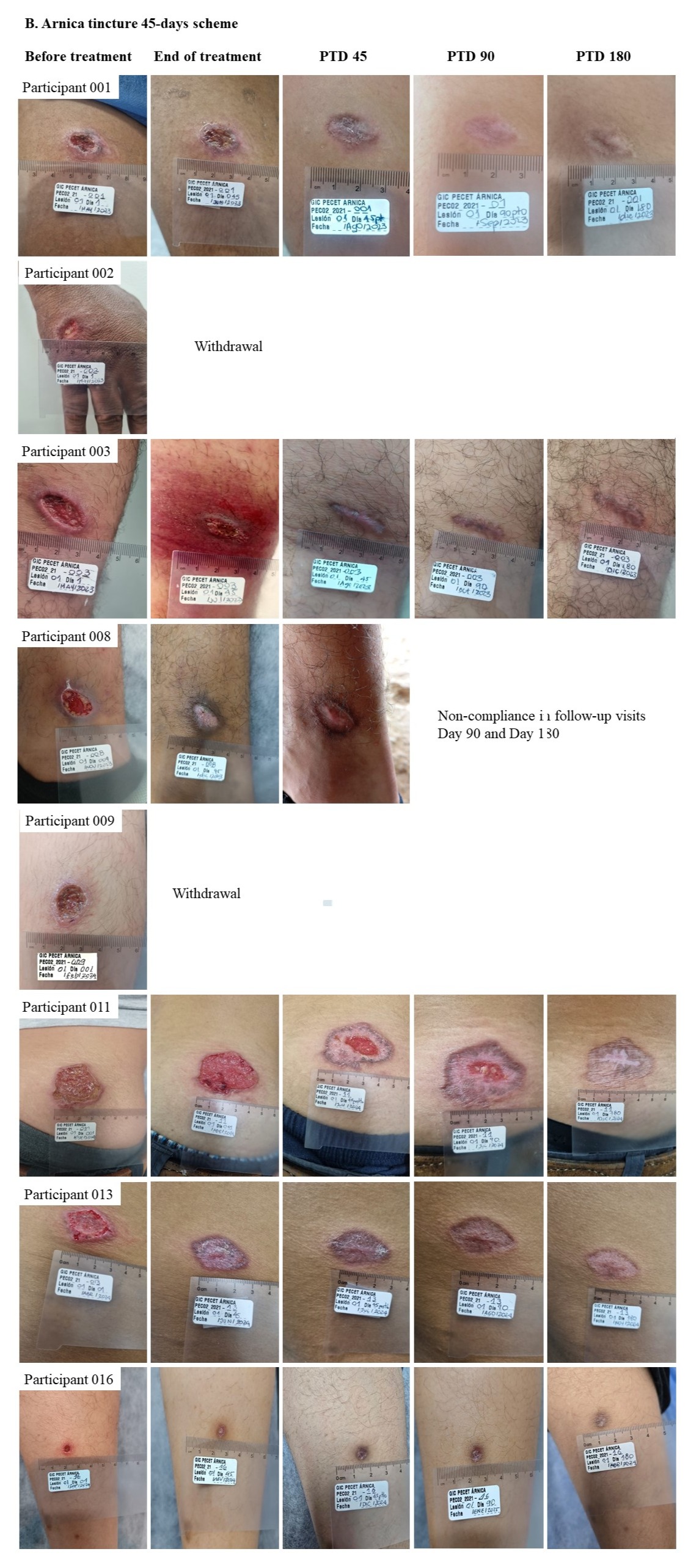
